# PFAS and phthalates in follicular fluid of a Nebraska in vitro fertilization (IVF) cohort

**DOI:** 10.64898/2026.09.21.26363443

**Authors:** Jabeen Taiba, Niranjana Balu, Ailenn C. Castillo, Rasha Barakat, Changyong Lee, Aimin Chen, Stephanie Gustin, Elizabeth Constance, John S. Davis, Jesse E. Bell, Kendra L. Clark

**Author notes:** **Corresponding author with contact information:** Jabeen Taiba, Department of Environmental, Agricultural, and Occupational Health, College of Public Health, University of Nebraska Medical Center, Omaha, NE, USA. Co-senior authors (Jesse E. Bell, Kendra L. Clark).

## Abstract

Per- and polyfluoroalkyl substances (PFAS) and phthalate metabolites remain understudied in follicular fluid of women undergoing assisted reproduction. We characterized these chemicals in previously collected follicular fluid from patients receiving fertility care at a Nebraska clinic (August 2022-June 2025) and linked exposure data with demographics and clinical characteristics from health records. Spearman correlations and nonparametric tests evaluated PFAS-phthalate relationships and exposure differences by age, race, BMI, and urbanicity. Seven PFAS and four phthalate metabolites were detected in at least half of participants. PFOS and PFHxS concentrations were higher in younger women, and mono-isobutyl phthalate (MiBP) concentrations were higher in younger and non-White women.

PFAS-phthalate correlations were weak and did not vary by urbanicity. These results characterize follicular fluid exposure patterns in a Midwestern cohort.

## 1. INTRODUCTION

Exposure to endocrine-disrupting chemicals (EDCs), per-and polyfluoroalkyl substances (PFAS) and phthalates may impact female fertility^1^. These compounds can cross the blood-follicle barrier and accumulate in the ovarian microenvironment of an oocyte^2^. Despite this, characterization of PFAS and phthalate exposure in human follicular fluid (FF) remains limited^3-7^. We quantified concentrations of legacy, short-chain, and ultra-short-chain) PFAS along with phthalates in the FF of women receiving fertility care in Nebraska, U.S., to characterize exposure patterns, assess exposure disparities within the cohort, and compare concentrations with those reported in other IVF cohorts.

## 2. METHODS

In this retrospective cohort study, previously collected follicular fluid samples from 139 patients undergoing IVF treatment from the Heartland Center for Reproductive Medicine (Omaha, Nebraska) between August 2022 to June 2025 (IRB#0147-22-EX) were utilized.

Samples were collected during transvaginal ultrasound-guided follicular aspiration approximately 36 h after ovulatory trigger administration, centrifuged to remove cellular components and stored at ™80°C until PFAS/phthalate analysis. PFAS were measured at Eurofins Environmental Testing Sacramento (California) and University of Nebraska Water Science labs. Phthalates were measured at NSF International (Ann Arbor, Michigan). Exposure measurement assays are described in the supplemental material. Analytes, acronyms, and limits of detection (LOD) are provided in Table S1. Analytes detected in >50% of the study population were included in further analysis. PFAS concentrations <LOD were imputed using left-truncated imputation method, whereas machine read values were used for phthalates.

Demographics and clinical characteristics, including age, race, body mass index (BMI), zip-code (aggregated as Rural/Urban), oocytes retrieved, dominant follicles, blastocyst progression, and final infertility diagnoses, were retrieved from the electronic health records. Exposure summary statistics [median, interquartile range (IQR; 25^th^ and 75^th^ percentiles] were calculated and compared with published studies reporting FF measures^3-7^. Spearman correlations quantified potential co-exposures among PFAS/phthalate analytes. Exposure disparities were assessed by comparing mean analyte concentrations (log-2 scale) across age, race, BMI, and urbanicity groups using non-parametric Kruskal Wallis and Wilcoxon signed-rank tests.

## 3. RESULTS

Overall, the mean age of participants was 34.4 years, and the mean BMI was 29.9 kg/m^2^ (Table 1). Most participants were White (111/139, 80%) and resided in urban areas (103/139, 74%). Demographic characteristics were similar across infertility diagnosis groups; however, clinical characteristics differed significantly. Participants with male factor of infertility had significantly more dominant follicles and oocytes retrieved, whereas a greater proportion of those with structural infertility (9/25, 36%) did not achieve clinical pregnancy.

**Table 1.** Participant characteristics by infertility diagnosis (n=139)

| Variable | Overall<br>(n=139) | Infertility diagnosis group |  |  |  | P-value |
| --- | --- | --- | --- | --- | --- | --- |
|  |  | Ovarian/<br>Hormonal<br>(n=67) | Structural<br>(n=25) | Other<br>(n=32) | Male factor<br>(n=15) |  |
| **<u>Demographics</u>** |  |  |  |  |  |  |
| Age (years) | 34.42 ± 5.37 | 35.57 ± 5.72 | 34.04 ± 4.29 | 32.94 ± 5.51 | 33.07 ± 4.22 | 0.054<sup>a</sup> |
| BMI (kg/m<sup>2</sup>) | 29.91 ± 6.76 | 30.26 ± 6.97 | 30.60 ± 7.64 | 28.22 ± 6.37 | 30.77 ± 4.79 | 0.271<sup>a</sup> |
| Race |  |  |  |  |  | 0.186<sup>b</sup> |
| White | 99 (80%) | 45 (80%) | 16 (73%) | 24 (77%) | 14 (100%) |  |
| Other | 24 (20%) | 11 (20%) | 6 (27%) | 7 (23%) | 0 (0%) |  |
| Residence (rural/urban) |  |  |  |  |  |  |
| Rural | 34 (26%) | 16 (25%) | 5 (21%) | 10 (33%) | 3 (20%) | 0.717<sup>b</sup> |
| Urban | 99 (74%) | 48 (75%) | 19 (79%) | 20 (67%) | 12 (80%) |  |
| **<u>Clinical characteristics</u>** |  |  |  |  |  |  |
| # oocytes retrieved | 16.30 ± 9.78 | 14.76 ± 11.09 | 16.80 ± 6.42 | 17.97 ± 10.03 | 18.80 ± 6.79 | 0.029<sup>a</sup> |
| # dominant follicles | 14.99 ± 8.52 | 12.84 ± 8.65 | 16.04 ± 8.49 | 16.59 ± 7.07 | 19.40 ± 8.86 | 0.004<sup>a</sup> |
| # progressed to blastocyst | 5.62 ± 5.22 | 5.18 ± 5.69 | 5.58 ± 3.98 | 5.58 ± 5.03 | 7.67 ± 5.14 | 0.152<sup>a</sup> |
| Insemination method |  |  |  |  |  |  |
| Conventional | 1 (1%) | 1 (1%) | 0 (0%) | 0 (0%) | 0 (0%) | 0.002<sup>b</sup> |
| ICSI | 128 (92%) | 65 (97%) | 24 (96%) | 24 (75%) | 15 (100%) |  |
| Other | 10 (7%) | 1 (1%) | 1 (4%) | 8 (25%) | 0 (0%) |  |
| Pregnant |  |  |  |  |  | <0.001<sup>b</sup> |
| Yes | 81 (58%) | 41 (61%) | 14 (56%) | 13 (41%) | 13 (87%) |  |
| No | 36 (26%) | 21 (31%) | 9 (36%) | 5 (16%) | 1 (7%) |  |
| Other | 22 (16%) | 5 (7%) | 2 (8%) | 14 (44%) | 1 (7%) |  |
| Live birth |  |  |  |  |  | 0.09<sup>b</sup> |
| Yes | 36 (26%) | 17 (25%) | 5 (20%) | 7 (22%) | 7 (47%) |  |
| No | 34 (24%) | 20 (30%) | 8 (32%) | 3 (9%) | 3 (20%) |  |
| Other | 69 (50%) | 30 (45%) | 12 (48%) | 22 (69%) | 5 (33%) |  |

Diagnoses were grouped as Male factor, Ovarian/Hormonal (diminished ovarian reserve, PCOS/PCO, advanced maternal age, ovarian dysfunction, hypothyroidism, hyperprolactinemia, Turner syndrome), Structural (tubal, endometriosis, adenomyosis, uterine/cervical factor, leiomyoma, recurrent pregnancy loss), or Other (unexplained, genetic, donor, fertility preservation, non-infertility). Multifactorial cases were assigned to a single group by a predefined clinical-priority hierarchy prioritizing ovarian/hormonal over structural and male factors. Race was categorized as White and Other (Black, Asian, Hispanic, Indian and Unknown). <sup>a</sup>Kruskal-Wallis test and <sup>b</sup>Fisher's exact.

Among the exposure analytes, 19 of 45 PFAS compounds and 12 of 14 phthalates were detected (Figure S1). Seven PFAS (perfluorocarboxylic acids and perfluorosulfonic acids) and four phthalates (low and high molecular weight) were detected in at least 50% of participants, and 62% had detectable concentrations of both PFAS and phthalates. Overall, PFAS concentrations were lower than those reported in IVF cohorts from Iowa^3^ (2019-2023; n=41), North Carolina^4^ (2024-2025; n=86), and Georgia^7^ (2018-2020; n=82) (Figure 1). By contrast, miBP concentrations were higher than those reported in a Danish cohort^5^ (2015-2017; n=111). PFAS showed moderate-to-strong correlations (r∼0.43–0.99), particularly among PFOS, PFOA, and PFHxS, consistent with shared exposure sources. Conversely, correlations between PFAS and phthalates (miBP, mCNP) were weak (r∼0.16–0.24). PFOS and PFHxS concentrations were significantly higher among women aged 19-37 years than those over 37 years, while miBP concentrations were higher in younger (19-32 years) and non-White women.

**Figure 1.**
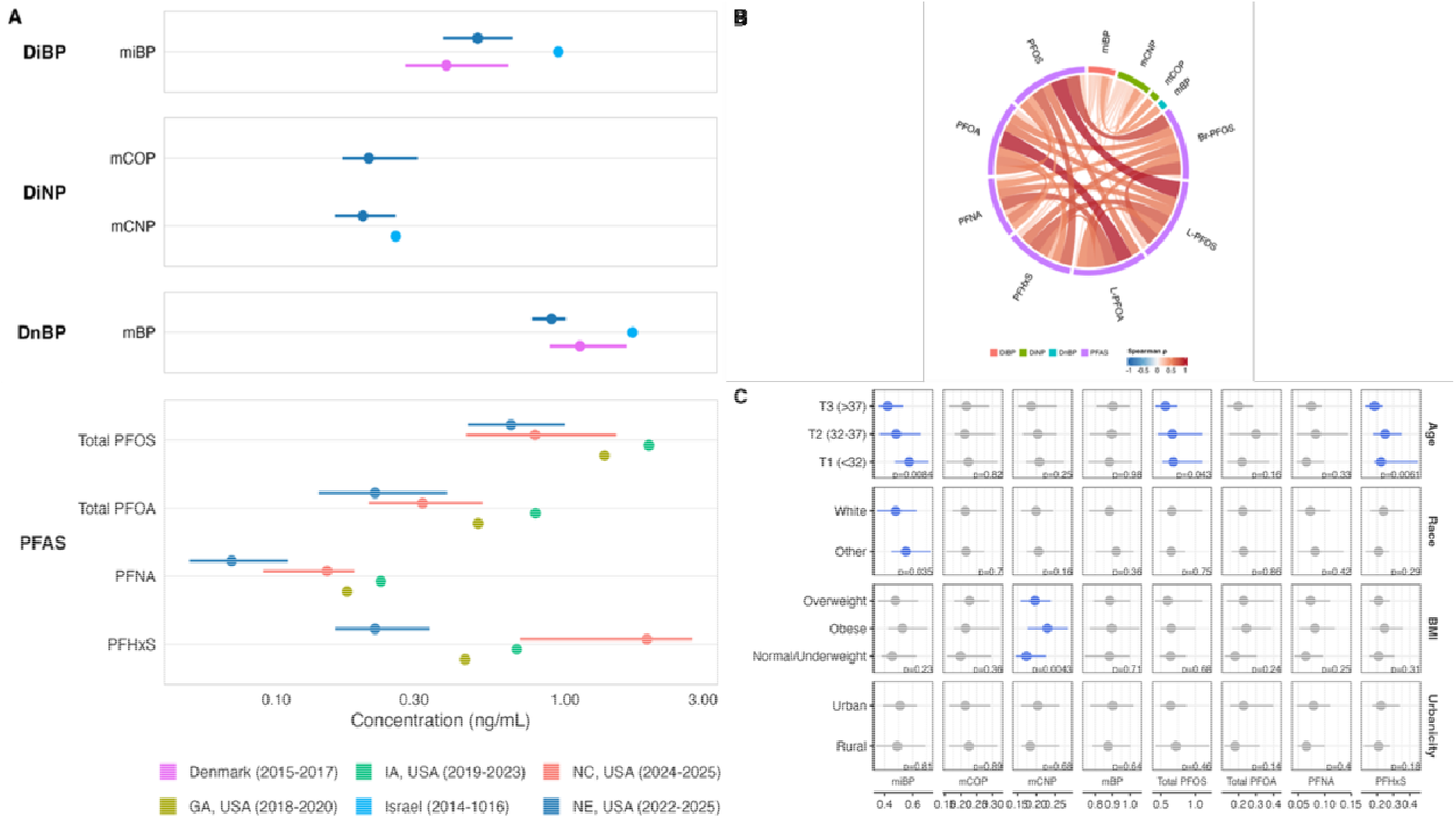
PFAS/Phthalate characterization. A-Comparision of exposure concentrations with other cohorts. B-Correlation between exposure analytes. C-Exposure differences by age, race, BMI and Urbanicity. Summary statistics for biomarkers detected in at least 25% study participants were added in Table S1.

## 4. DISCUSSION

We detected both legacy and emerging PFAS compounds as well as low- and high-molecular weight phthalates in FF samples collected from women in the Midwestern-US. PFAS and phthalates concentrations were generally lower than those reported in other populations, potentially reflecting differences in demographic characteristics, dietary patterns, and proximity to industrial sources. PFOS and PFHxS concentrations were significantly higher among women aged 19-37 years than those older than 37 years, a finding that contrasts with biological plausibility, as reproductive elimination through menstruation/parity can reduce PFAS body burden^8^. Among phthalate biomarkers, miBP concentrations were higher in younger (19–32 years) and non-White women, consistent with differences in personal care product use across age and race/ethnicity^9^. mCNP concentrations were higher in obese than overweight or underweight women, possibly reflecting greater dietary exposure, as DiNP (the parent compound of mCNP) is used primarily in food packaging and associated with consumption of ultra-processed foods^10^. A limitation of our study was the lack of comprehensive demographic data to further explain determinants of these exposures. Future studies should evaluate associations between PFAS/phthalate biomarkers in FF and IVF outcomes and more fully characterize exposure disparities by age and race/ethnicity in larger and more racially diverse populations.

## Data Availability

All data produced in the present study are available upon reasonable request to the authors

## CRediT Authorship Contribution Statement

**Jabeen Taiba:** Writing – original draft, Visualization, Methodology, Formal analysis, Data curation, Conceptualization, Funding acquisition. **Niranjana Balu:** Clinical data acquisition, Writing – review & editing. **Ailenn Castillo:** Biospecimen collection, Writing – review & editing. **Rasha Barakat:** Biospecimen processing, Writing – review & editing. **Changyong Lee:** Biospecimen processing, Writing – review & editing. **Aimin Chen**: Validation, Writing – review & editing. **Stephanie Gustin:** Project administration, Biospecimen collection, Writing – review & editing. **Elizabeth Constance:** Project administration, Biospecimen collection, Writing – review & editing. **John S. Davis:** Project administration, Writing – review & editing. **Jesse E. Bell:** Funding acquisition, Supervision, Validation, Project administration, Writing – review & editing. **Kendra Clark:** Writing – review & editing, Validation, Supervision, Methodology, Funding acquisition, Conceptualization.

## References

1. Panagiotou, E.M., Ojasalo, V. & Damdimopoulou, P. Phthalates, ovarian function and fertility in adulthood. Best Pract Res Clin Endocrinol Metab 35, 101552 (2021).

2. Bjorvang, R.D., et al. Follicular fluid and blood levels of persistent organic pollutants and reproductive outcomes among women undergoing assisted reproductive technologies. Environ Res 208, 112626 (2022).

3. Good, S.L., et al. Molecular structure, isomerism, and concentration as determinants of PFAS transfer from plasma to ovarian follicular fluid. Sci Total Environ 1030, 181773 (2026).

4. Boney, S.S., et al. First evidence of legacy and emerging per- and polyfluoroalkyl substances (PFAS) in the follicular fluid of a cohort of North Carolina in vitro fertilization (IVF) patients. Reprod Toxicol 139, 109102 (2026).

5. Beck, A.L., et al. Ovarian follicular fluid levels of phthalates and benzophenones in relation to fertility outcomes. Environ Int 183, 108383 (2024).

6. Hoffmann-Dishon, N., et al. Endocrine-disrupting chemical concentrations in follicular fluid and follicular reproductive hormone levels. J Assist Reprod Genet 41, 1637–1642 (2024).

7. Young, A.S., et al. Integrated chemical exposome-metabolome profiling of follicular fluid and associations with fertility outcomes during assisted reproduction. Environ Int 203, 109787 (2025).

8. Wong, F., MacLeod, M., Mueller, J.F. & Cousins, I.T. Enhanced elimination of perfluorooctane sulfonic acid by menstruating women: evidence from population-based pharmacokinetic modeling. Environ Sci Technol 48, 8807–8814 (2014).

9. James-Todd, T.M., et al. Racial and ethnic variations in phthalate metabolite concentration changes across full-term pregnancies. J Expo Sci Environ Epidemiol 27, 160–166 (2017).

10. Baker, B.H., et al. Ultra-processed and fast food consumption, exposure to phthalates during pregnancy, and socioeconomic disparities in phthalate exposures. Environ Int 183, 108427 (2024).

