## Supplemental File for "PFAS and phthalates in follicular fluid of a Nebraska in vitro fertilization (IVF) cohort"

Contents (7 pages): 1 table, 1 figure

**Table of contents:**

Table S1. Biomarker limit of detection and detection frequency ……………………….……1

Figure S1. Unique detection frequency of PFAS/Phthalate biomarkers …………………… 6

PFAS and Phthalate biomarker assays ……………………………………………………….. 7

| **Table S1. Biomarker limit of detection and detection frequency (n=139)** | | | | | |  |
| --- | --- | --- | --- | --- | --- | --- |
| **Class** | **Subgroup/parent compound** | **Analyte** | **Abbreviation** | **MDL (ng/mL)** | **Percent detection** | **Median (IQR)** |
| **Legacy PFAS (C8+ PFCAs; C7+ PFSAs)** | Perfluorocarboxylic acids (PFCAs) | L-Perfluorooctanoic acid | *L-PFOA* | 0.14 | 76.6 | 0.22 (0.14, 0.36) |
|  |  | Br-Perfluorooctanoic acid | *Br-PFOA* | 0.14 | 0 |  |
|  |  | Total perfluorooctanoic acid | *PFOA* | 0.14 | 76.1 | 0.22 (0.14, 0.39) |
|  |  | Perfluorononanoic acid | *PFNA* | 0.050 | 68.8 | 0.07 (0.05, 0.11) |
|  |  | Perfluorodecanoic acid | *PFDA* | 0.050 | 10.2 |  |
|  |  | Perfluoroundecanoic acid | *PFUnA* | 0.050 | 7.8 |  |
|  |  | Perfluorododecanoic acid | *PFDoA* | 0.050 | 0 |  |
|  |  | Perfluorotridecanoic acid | *PFTrDA* | 0.050 | 1.6 |  |
|  |  | Perfluorotetradecanoic acid | *PFTeDA* | 0.050 | 0 |  |
|  |  | Perfluoro-n-hexadecanoic acid | *PFHxDA* | 0.050 | 0 |  |
|  | Perfluorosulfonic acids (PFSAs) | Perfluoroheptanesulfonic acid | *PFHpS* | 0.10 | 0 |  |
|  |  | L-Perfluorooctanesulfonic acid | *L-PFOS* | 0.050 | 100 | 0.51 (0.37, 0.76) |
|  |  | Br-Perfluorooctanesulfonic acid | *Br-PFOS* | 0.050 | 89.8 | 0.12 (0.08, 0.17) |
|  |  | Total perfluorooctanesulfonic acid | *PFOS* | 0.050 | 100 | 0.65 (0.46, 1.00) |
|  |  | Perfluorononanesulfonic acid | *PFNS* | 0.050 | 0 |  |
|  |  | Perfluorodecanesulfonic acid | *PFDS* | 0.050 | 1.6 |  |
|  |  | Perfluorododecanesulfonic acid | *PFDoS* | 0.050 | 0 |  |
|  | Sulfonamides and precursors | Perfluorooctanesulfonamide | *PFOSA* | 0.050 | 0 |  |
|  |  | N-methyl perfluorooctanesulfonamido acetic acid | *NMeFOSAA* | 0.050 | 5.8 |  |
|  |  | N-ethyl perfluorooctanesulfonamido acetic acid | *NEtFOSAA* | 0.050 | 4.4 |  |
|  | Fluorotelomer compounds | 7:3 Fluorotelomer carboxylic acid | *7:3 FTCA* | 0.050 | 0 |  |
|  |  | 8:2 Fluorotelomer carboxylic acid | *8:2 FTCA* | 0.056 | 0 |  |
|  |  | 8:2 Fluorotelomer sulfonic acid | *8:2 FTS* | 0.050 | 0 |  |
|  |  | 10:2 Fluorotelomer sulfonic acid | *10:2 FTS* | 0.050 | 0 |  |
|  |  | 8:2 Fluorotelomer unsaturated carboxylic acid | *8:2 FTUCA* | 0.050 | 0 |  |
| **Short-chain PFAS (C4-C7 PFCAs; C4-C6 PFSAs)** | Perfluorocarboxylic acids (PFCAs) | Perfluorohexanoic acid | *PFHxA* | 0.10 | 0 |  |
|  |  | Perfluoroheptanoic acid | *PFHpA* | 0.050 | 21.6 |  |
|  | Perfluorosulfonic acids (PFSAs) | Perfluoropentanesulfonic acid | *PFPeS* | 0.050 | 0.8 |  |
|  |  | Perfluorohexanesulfonic acid | *PFHxS* | 0.050 | 97.8 | 0.22 (0.16, 0.34) |
|  | Fluorotelomer compounds | 5:3 Fluorotelomer carboxylic acid | *5:3 FTCA* | 0.096 | 0 |  |
|  |  | 6:2 Fluorotelomer carboxylic acid | *6:2 FTCA* | 0.069 | 0 |  |
|  |  | 4:2 Fluorotelomer sulfonic acid | *4:2 FTS* | 0.050 | 0 |  |
|  |  | 6:2 Fluorotelomer unsaturated carboxylic acid | *6:2 FTUCA* | 0.067 | 0 |  |
| **Ultra-short-chain and novel/replacement PFAS (≤C3 and ether-PFAS)** | Ether carboxylic acids | Hexafluoropropylene oxide dimer acid (GenX) | *HFPO-DA* | 0.10 | 0.8 |  |
|  |  | 4,8-Dioxa-3H-perfluorononanoic acid | *ADONA* | 1.0 | 0 |  |
|  |  | Perfluoro(2-methoxyacetic acid) | *PFMBA* | 0.050 | 0 |  |
|  |  | Perfluoro-2-(ethoxy)ethanesulfonic acid | *PFEESA* | 0.050 | 0 |  |
|  |  | Perfluoro-3,5,7,9,11-pentaoxadodecanoic acid | *PFO5DA* | 0.10 | 0 |  |
|  |  | Perfluoropolyether carboxylic acid | *PFPE-1* | 0.12 | 0 |  |
|  |  | Nonafluoro-3,6-dioxaheptanoic acid | *NFDHA* | 0.050 | 0 |  |
|  | Ether sulfonic acids | 9-Chlorohexadecafluoro-3-oxanone-1-sulfonic acid | *9Cl-PF3ONS* | 0.050 | 1.5 |  |
|  |  | 11-Chloroeicosafluoro-3-oxaundecane-1-sulfonic acid | *11Cl-PF3OUdS* | 0.050 | 0 |  |
|  |  | Perfluoroethylcyclohexanesulfonic acid | *PFECHS* | 0.050 | 0 |  |
|  | Short-chain perfluorosulfonate | Perfluorobutanesulfonic acid | *PFBS* | 0.20 | 0.8 |  |
|  | Hydrogenated PFAS | Hydrogenated perfluorosulfonic acid | *Hydro-PS Acid* | 0.050 | 0 |  |
| **Phthalates** | | | | | |  |
| **High-molecular-weight (HMW) phthalates** | Di(2-ethylhexyl) phthalate (DEHP) | Mono(2-ethylhexyl) phthalate | *mEHP* | 1.0 | 11.5 |  |
|  |  | Mono(2-ethyl-5-hydroxyhexyl) phthalate | *mEHHP* | 0.1 | 1.4 |  |
|  |  | Mono(2-ethyl-5-oxohexyl) phthalate | *mEOHP* | 0.1 | 0.72 |  |
|  |  | Mono(2-ethyl-5-carboxypentyl) phthalate | *mECPP* | 0.2 | 28.1 | <LOD (<LOD, 0.21) |
|  |  | Mono(3-carboxypropyl) phthalate | *mCPP* | 0.2 | 0 |  |
|  | Benzyl butyl phthalate (BBzP) | Mono-benzyl phthalate | *mBzP* | 0.2 | 18.7 |  |
|  | Di-isononyl phthalate (DiNP) | Mono-isononyl phthalate | *mNP* | 0.5 | 0.72 |  |
|  |  | Mono-carboxy-isononyl phthalate | *mCNP* | 0.2 | 51.1 | 0.20 (<LOD, 0.26) |
|  | Di-isooctyl phthalate (DiOP) | Mono-carboxy-isooctyl phthalate | *mCOP* | 0.2 | 56.8 | 0.21 (<LOD, 0.31) |
|  | Di(isononyl) cyclohexane-1,2-dicarboxylate (DINCH) | Cyclohexane-1,2-dicarboxylic acid mono-carboxyisooctyl ester | *CX-MINCH* | 0.2 | 2.2 |  |
|  |  | Cyclohexane-1,2-dicarboxylic acid mono-hydroxyisononyl ester | *OH-MINCH* | 0.2 | 0 |  |
| **Low-molecular-weight (LMW) phthalates** | Di-n-butyl phthalate (DnBP) | Mono-n-butyl phthalate | *mBP* | 0.5 | 99.3 | 0.90 (0.77, 1.01) |
|  | Di-isobutyl phthalate (DiBP) | Mono-isobutyl phthalate | *miBP* | 0.1 | 100 | 0.50 (0.38, 0.66) |
|  | Diethyl phthalate (DEP) | Mono-ethyl phthalate | *mEP* | 1.0 | 10.1 |  |
| PFCAs, perfluorocarboxylic acids; PFSAs, perfluorosulfonic acids; MDL, method detection limit. Legacy PFAS are defined as perfluorocarboxylic acids with ≥ 8 carbons and perfluorosulfonic acids with ≥ 7 carbons; short-chain PFAS as perfluorocarboxylic acids with 4–7 carbons and perfluorosulfonic acids with 4–6 carbons; ultra-short-chain and novel/replacement PFAS include ≤ 3-carbon and ether-linked perfluorinated compounds. HMW, high molecular weight; LMW, low molecular weight; MDL, method detection limit. High-molecular-weight phthalates have a parent-diester alcohol side chain of ≥ 6 carbons; low-molecular-weight phthalates have a side chain of < 6 carbons. DINCH is a non-phthalate plasticizer included here as a phthalate substitute. Median and IQR (25^th^ and 75^th^ percentile) values are reported for biomarkers detected in at least 25% of the study population. | | | | | |  |

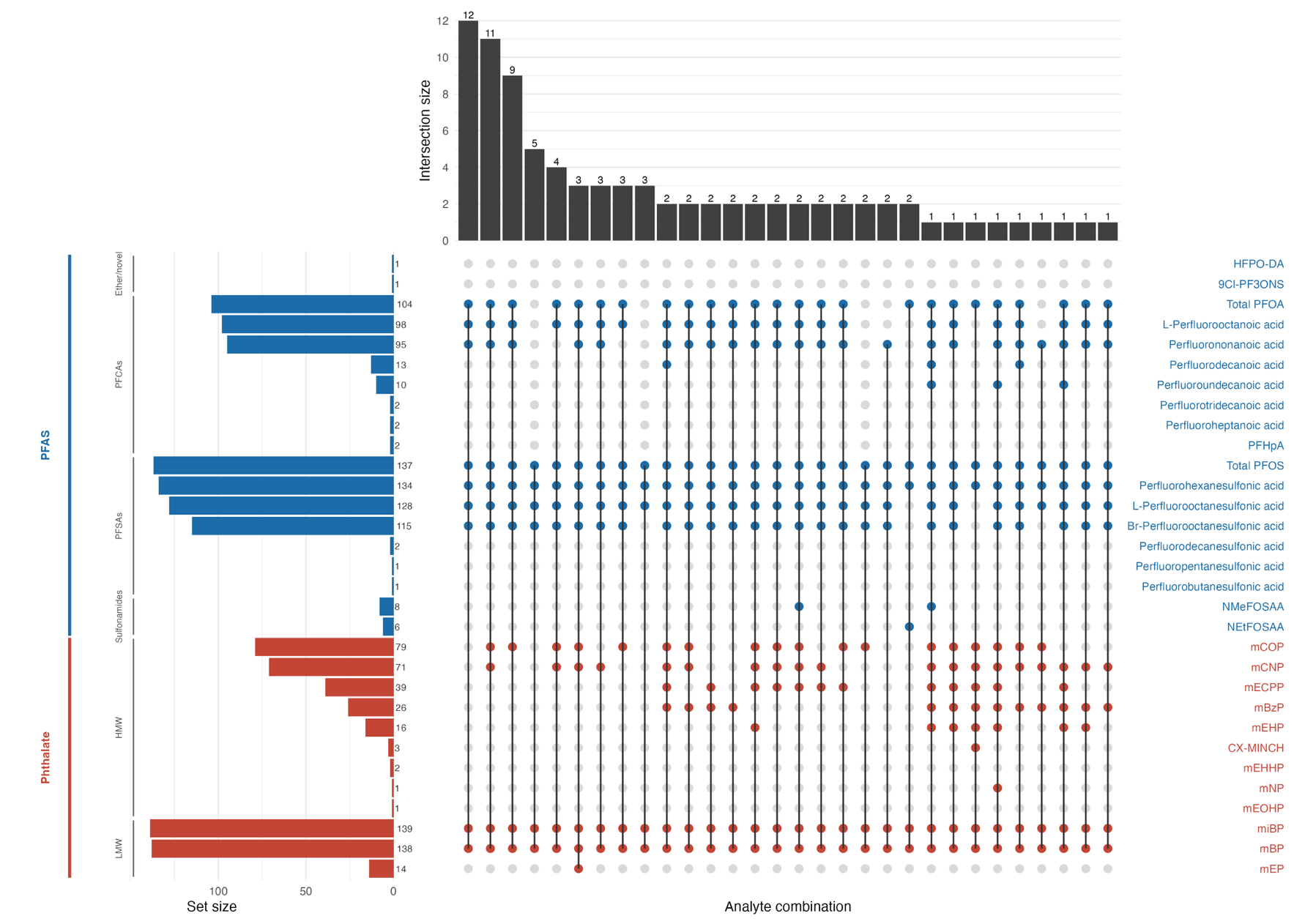

Figure S1. Unique detection frequency of PFAS/Phthalate biomarkers. Exposure biomarkers listed on right side and color coded into two groups, Blue=PFAS biomarkers & Red=Phthalate biomarkers. PFAS/Phthalate biomarker sub-groups were further stratified in the left. Bar plots and labels on the y-axis represents number of participants with corresponding biomarker concentrations above the LOD. Bar plots/labels on the x-axis and the dot matrix representing unique combination of biomarkers visualize number of participants with unique biomarker detection patterns.

PFAS and Phthalate biomarker assays: PFAS biomarkers were measured following the EPA method 537.1 (EPA/600/R-18/352) for Branched, Linear, and Total (B/L/T) PFAS. This method uses solid-phase extraction (SPE) and liquid chromatography-tandem mass spectrometry (MS/MS) for biomarker quantification. Phthalate biomarkers were measured {Good, 2026 #41;Boney, 2026 #5;Young, 2025 #7;Hoffmann-Dishon, 2024 #15;Beck, 2024 #40}using solid-phase extraction coupled to high-performance liquid chromatography-isotope dilution tandem mass spectrometry (Method No: 6306.03). Briefly, conjugated species of phthalate metabolites were hydrolyzed using β-glucuronidase/sulfatase, and compounds of interest were concentrated by online SPE, separated by reversed-phase high-performance liquid chromatography, and detected using atmospheric-pressure chemical ionization (APCI)-MS/MS.
